# Quantitative susceptibility mapping improves the detection of calcified carotid vessels by multi-contrast MRI using computed tomography angiography as the reference standard

**DOI:** 10.1101/2021.07.07.21260049

**Authors:** Thanh D. Nguyen, Yan Wen, Jingwen Du, Pascal Spincemaille, Yi Wang, Yang Qi, Hooman Kamel, Ajay Gupta

## Abstract

The objective of this study was to evaluate initial feasibility and improvement in the detection of calcified carotid arteries by including quantitative susceptibility mapping (QSM) in the carotid vessel wall multi-contrast MRI (mcMRI) protocol using CTA as the reference standard. In a pilot cohort of ten patients with significant carotid artery stenosis, calcified vessel detection by mcMRI achieved 64.7% sensitivity and 100% specificity. Adding QSM to mcMRI improved sensitivity to 100% while not affecting specificity.

## INTRODUCTION

Calcification is a common feature of carotid atherosclerotic plaques and may contribute to plaque stability^1^, thereby influencing a patient’s risk of stroke^2^. Accurate assessment of plaque calcification is therefore critically important in fully characterizing the risk profile of carotid atherosclerotic plaques and may help to identify those patients who would most benefit from invasive revascularization, versus those with lower risk who would benefit from noninvasive medical therapy. Currently, computed tomography angiography (CTA) is considered the gold standard imaging modality for the in vivo detection and quantification of plaque calcification^4^. While efficient and highly standardized, this technique requires harmful radiation and does not provide adequate characterization of other important plaque components such as intraplaque hemorrhage (IPH)^5^. Multi-contrast MRI (mcMRI) has the ability to fully characterize plaque composition without CTA limitations^4^. However, accurate interpretation of calcification on mcMRI, which is based on T1W and T2W hypointensity, can be difficult as hemosiderin-rich IPH, perivascular fat, muscle tissues, and blood vessels may also appear hypointense on the black blood fat-suppressed mcMRI images. Recently, quantitative susceptibility mapping (QSM) has been shown to provide a novel contrast for resolving this mcMRI ambiguity^6-8^, because calcification is a uniquely strong diamagnetic source in the body and is easily distinguishable from paramagnetic sources such as hemorrhage and fat. The objective of this pilot study was to evaluate the improvement in diagnostic accuracy for calcified vessel detection by adding QSM to mcMRI, using CTA as the reference standard.

## METHODS

Carotid QSM was evaluated in ten male patients (mean age, 66 years ± 5) who had hemodynamically significant (>50%) stenosis of at least one carotid artery and were undergoing 3T mcMRI and CTA for clinical purposes. The median follow-up interval between MRI and CTA scans was 3 days. The carotid mcMRI protocol was based on ASNR Vessel Wall Imaging Study Group recommendations^4^ and consisted of: 1) axial 3D TOF: TR/TE=20/3.6ms, flip angle (FA)=30°, bandwidth=250 Hz/pixel, voxel=0.63×0.63×2mm^3^, 4:03 min; 2) axial 2D black blood fat-suppressed T1W TSE: TR/TE=885/9.4ms, ETL=7, NEX=2, bandwidth=410 Hz/pixel, voxel=0.63×0.63×2mm^3^, 4:12 min; 3) axial 2D black blood fat-suppressed T2W TSE: TR/TE=4770/58 ms, ETL=12, NEX=3, bandwidth=410 Hz/pixel, voxel=0.63×0.63×2mm^3^, 6:12 min; 4) coronal 3D magnetization-prepared rapid gradient echo (MPRAGE): TR/TE/TI= 840/3.4/500ms, FA=15°, bandwidth=331 Hz/pixel, voxel size=0.63×0.63×0.83mm^3^, 3:44 min. QSM was obtained using a multi-echo 3D gradient echo sequence with the following parameters: TR/first TE/ΔTE=21/2.9/4.7ms, ETL=4, FA=10°, NEX=2, bandwidth=580 Hz/pixel, voxel=0.63×0.63×2mm^3^, 8:03 min. QSM images were reconstructed using the nonlinear preconditioned total field inversion algorithm using the MEDI toolbox^6^. A neuroradiologist with 13 years of carotid imaging experience independently reviewed CTA, mcMRI, and mcMRI+QSM images in three sessions (at least three weeks apart to minimize recall bias) to identify plaque calcification on a per vessel basis. Calcification was detected as a region with high attenuation on CTA (Hounsfield unit>130)^4^, hypointense signal (compared to the adjacent sternocleidomastoid muscle) on mcMRI, and strongly negative susceptibility (<-0.5 ppm) on QSM.

## RESULTS

Figure 1 shows an example of concordant depiction by mcMRI, QSM, and CTA of a heavily calcified plaque at the carotid bifurcation. Compared to mcMRI, the unambiguous image contrast between calcification and the surrounding tissues on QSM allows easy identification and improves diagnostic confidence. This benefit is further highlighted in Figure 2 and Supplement Figure S1, which show two examples of small calcification nodules from two other patients with subtle or ambiguous signal that could not be prospectively identified by the expert reader on mcMRI but well captured by QSM (approximately -2 and -1.5 ppm susceptibility, respectively) in excellent agreement with CTA. Out of 17 vessels with calcified plaques detected by CTA, mcMRI only captured 11 calcifications (64.7% sensitivity, 95% confidence interval 43.8%-85.6%). However, with the addition of QSM to mcMRI, all calcified vessels were identified (100% sensitivity). Both mcMRI and mcMRI+ QSM correctly identified the three non-calcified vessels (100% specificity).

**Figure 1.**
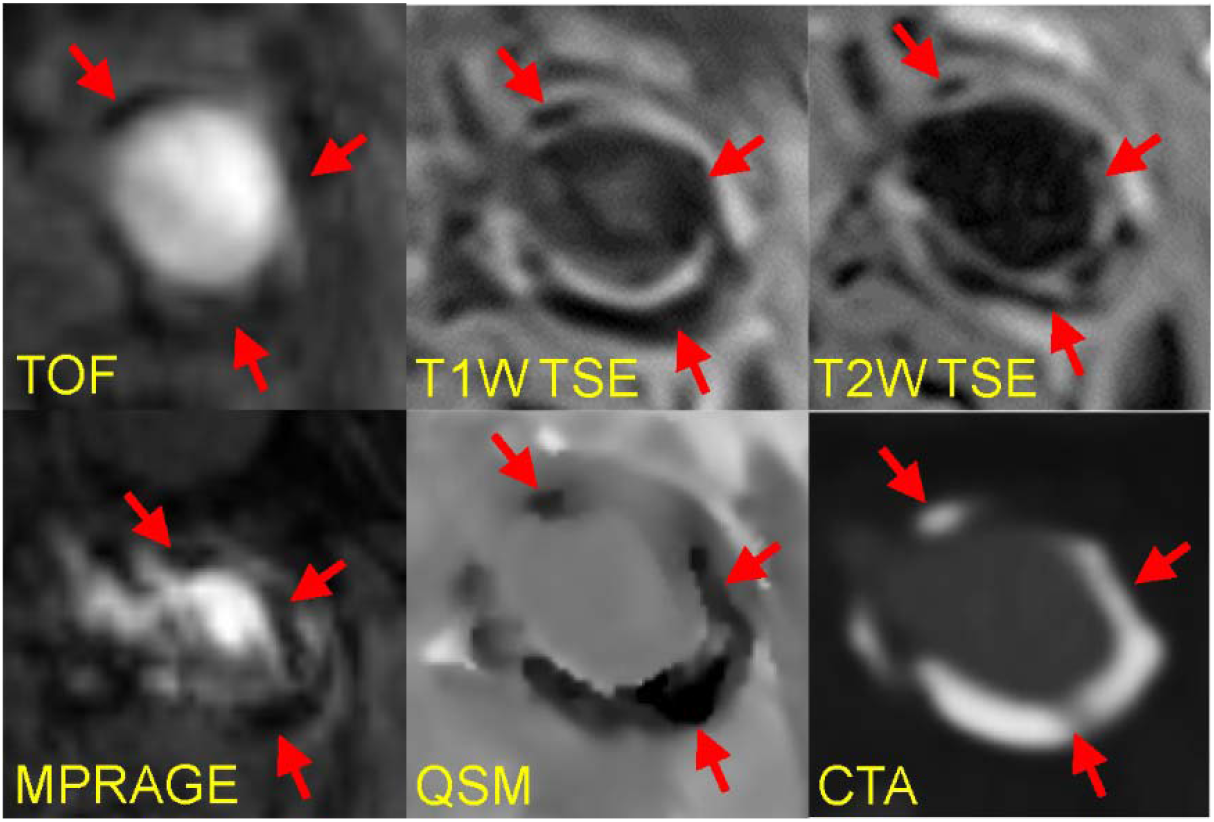
Example of concordant detection by traditional mcMRI, QSM, and CTA of a heavy concentric calcification near the left carotid bifurcation.

**Figure 2.**
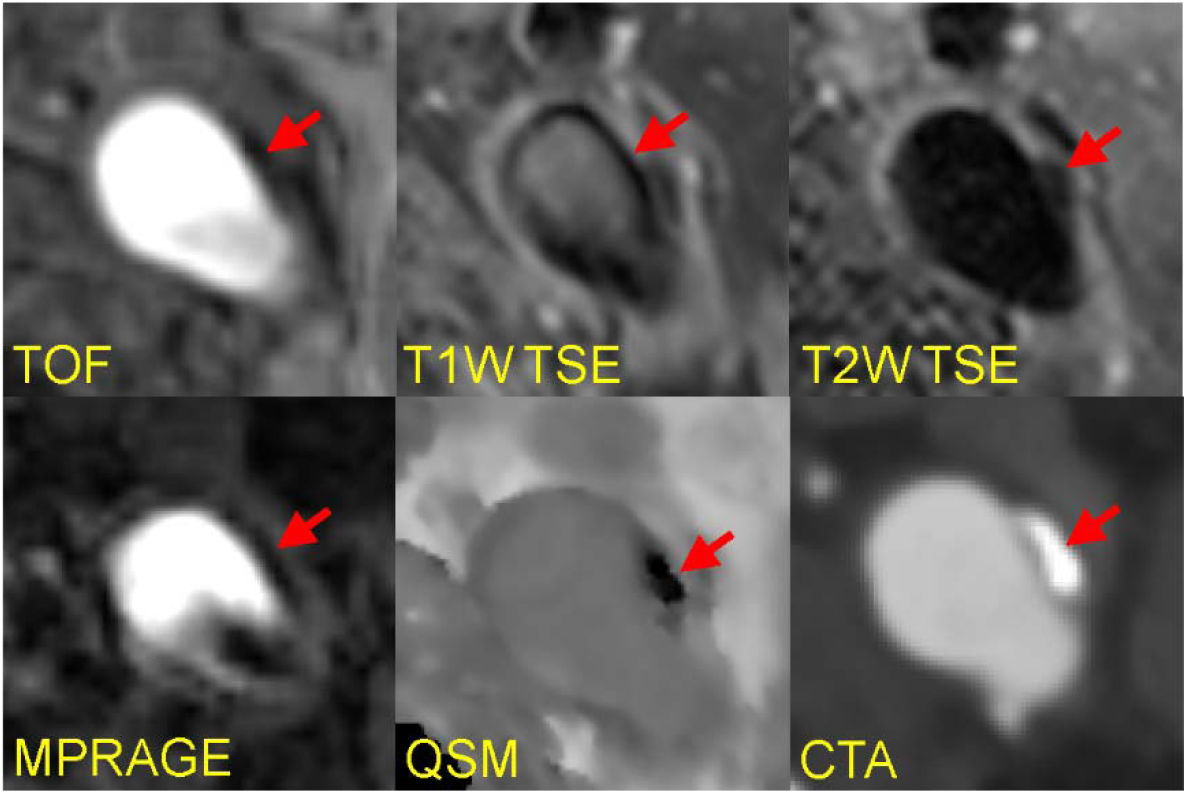
Example of a small calcified nodule in the left carotid artery that could not be prospectively identified on traditional mcMRI due to poor lesion contrast but was detected with high confidence by the expert reader on QSM in excellent agreement with CTA.

## DISCUSSION

We evaluated the benefit of adding QSM to the carotid mcMRI protocol using CTA as the in vivo reference standard for calcification detection and found that QSM improves the visual depiction of calcified plaques and substantially increases the detection sensitivity of mcMRI. QSM is known to provide accurate and sensitive detection of strong magnetic materials in the human body^9^, including highly paramagnetic iron in hemorrhage and highly diamagnetic calcification deposits. While our pilot study has a limited sample size, the promising results presented here show that QSM has the potential to become an instrumental part of noninvasive and quantitative carotid mcMRI for plaque characterization and warrants further evaluation in larger clinical studies.

## Data Availability

Data are available to interested researchers upon reasonable request.

---

**Figure S1.**
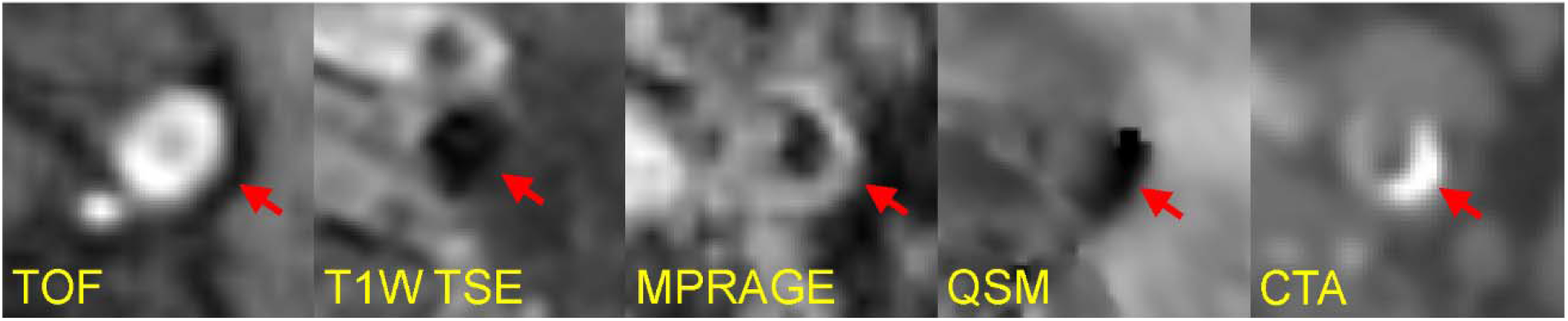
Example of a small calcified plaque in the internal carotid artery that was not detected by mcMRI but well visualized on QSM and confirmed by CTA.

